# Enhanced Adverse-Event Detection and Drug-Event Relation Extraction from Clinical Notes

**DOI:** 10.64898/2026.05.06.26352616

**Authors:** Omar Alharbi, Cathy H Wu, Chuming Chen, K Vijay-Shanker

**Affiliations:** Center for Bioinformatics and Computational Biology University of Delaware, Newark, DE, USA; Department of Computer and Information Sciences University of Delaware, Newark, DE, USA

**Keywords:** Adverse drug event, ADE, Adverse events, AE, clinical notes, pharmacovigilance, relation extraction

## Abstract

Adverse drug events (ADEs) are a significant source of preventable patient harm, yet many ADE signals remain buried in free-text clinical notes. Clinical notes often describe adverse events (AEs) in relation to drugs in two ways: whether a drug causes the AE (the AE is an ADE) or a drug is given to treat an AE (it is considered the Reason for drug treatment). In the N2C2 2018 benchmark, ADEs and Reasons are annotated as separate entity types, despite often being similar in both wording and clinical meaning. This shared similarity makes them difficult to distinguish during entity extraction, leading to errors in relation classification. Therefore, we propose a two-stage framework that first detects AEs as a unified event category and then classifies drug-event pairs into Drug-ADE, Drug-Reason, or No-Relation. In the end-to-end evaluation on the N2C2 2018 benchmark, our system achieves F1 scores of 0.93 for Drug-ADE and 0.94 for Drug-Reason, improving over previously reported end-to-end benchmarks of 0.48 for Drug-ADE and 0.59 for Drug-Reason. Overall, these results support a more precise task formulation in which AEs are detected broadly first, and the ADE vs Reason distinction is resolved at the relation layer. Furthermore, they motivate the development of AE-focused datasets annotated independently of drug linkage to enable more reliable end-to-end pharmacovigilance systems.

## I. Introduction

Adverse Drug Events (ADEs) are any harmful or unpleasant reactions that result from the use of medicinal products [1]. In 2022, more than 1.25 million serious adverse drug events were reported to the FDA’s FAERS system, including nearly 175,000 deaths [2]. ADEs lead to over 1.5 million emergency department visits annually in the U.S., with nearly 500,000 cases resulting in hospitalization [3]. Several adverse events can occur to patients during their hospital treatment. Those events vary by type and are primarily defined in relation to what has caused them. Accurately detecting ADEs in clinical notes is imperative for enhancing patient safety and supporting pharmacovigilance.

Like ADEs, we can say that Adverse Events (AEs) are any undesirable experiences or clinical outcomes that occur during a patient’s care and may or may not be causally related to a therapeutic intervention. When examined in relation to medications, AEs are typically classified into two categories: Adverse Drug Events (ADEs), in which the drug causes harm or unintended effects, or “Reasons,” in which the drug is administered to treat an existing AE or underlying condition. Much of the evidence needed to identify and distinguish these cases is recorded in Electronic Health Records (EHRs), particularly within clinicians’ documentation [4]. While EHRs also contain structured elements such as medications and laboratory results, AEs are often described in narrative clinical notes that capture nuanced observations, context, and clinical reasoning, which are important for pharmacovigilance and related prediction tasks [5]. However, the free-text nature of these notes, along with their linguistic and formatting variability, creates significant challenges for automated AE detection and analysis.

Information extraction (IE) from clinical notes has become an essential task in healthcare. Due to the nature and format of the data in these notes, data scientists rely on natural language processing (NLP) techniques to extract information. The extraction of drug information from clinical notes is a crucial task that facilitates a deeper understanding of patient care practices and medication management, and reduces medical errors [6]. Accurate extraction of drug-related entities enables identification of drug names, dosages, routes, and frequencies, which are essential for patient safety [7]. Furthermore, detecting ADEs requires accurately identifying both the drug and the related adverse event, making the Named Entity Recognition (NER) task crucial for extracting and linking these mentions. Relation extraction (RE) then determines whether the AE is an ADE or a Reason.

Detection of drug-related adverse events (i.e., ADE as well as Reason) remains a persistent challenge for automated NLP systems, as current methodologies have limitations that reduce their effectiveness in real-world documentation where relevant evidence is often buried in narrative text. The goal of this work is to develop an end-to-end NLP framework that more robustly identifies drug-related adverse events from clinical notes and determines their relationships to medications. Using this approach, we significantly improve end-to-end Drug-ADE extraction performance, increasing F1 score from 0.48 to 0.93, demonstrating a substantial improvement over the prior state-of-the-art. The main contributions of this study are:

1. **Preliminary analysis:** Identified a key limitation of treating ADE and Reason as separate concept types in the NER stage before relation classification.
2. **Reformulated the task:** Proposed a unified adverse event (AE) concept that generalizes over both ADE and Reason at the mention level.
3. **AE detection methods:** Developed methods to detect AE mentions in clinical notes to support downstream relation modeling.
4. **End-to-end system:** Built an end-to-end drug-event extraction system that significantly outperforms prior results on the n2c2 2018 benchmark.

## II. Related work

The 2018 N2C2 shared task (“Track 2: Adverse Drug Events and Medication Extraction”) provided a standardized evaluation framework for extracting medication-related concepts and their relations from narrative discharge summaries [8]. Participants were tasked with three subtasks: Concept Extraction, Relation Classification, and End-to-End system development, using a corpus of 505 annotated clinical notes from the MIMIC-III database. Each note had been manually annotated for eight concept types (including Drug, Strength, Form, Dosage, Frequency, Route, Duration, Reason, and ADE) and nine relation categories linking those concepts to drug mentions in raw clinical text. For evaluation, systems were assessed using a lenient micro-averaged F1. While the overall top lenient F1 reached 0.9418, ADE concepts proved especially challenging: even among the best teams, ADE recognition lagged far behind other categories, achieving only about 0.58 lenient F1.

The MADE 1.0 initiative established one of the first comprehensive benchmarks for NLP in EHRs for drug safety surveillance [9]. Its corpus contains 1,089 fully de-identified longitudinal notes drawn from 21 cancer patients at UMass Memorial, spanning discharge summaries, consult reports, and related documents. Moreover, it uses the same evaluation structure as the N2C2 2018 Track 2 ADE challenge, with the same three shared tasks released: concept extraction, relation classification, and end-to-end evaluation. Although the top-performing model, a bidirectional LSTM-CRF model, achieved a strict micro-F1 of 0.82 overall, performance on the most safety-critical concepts was much lower: ADE recognition peaked at F1 of 0.61, and indication recognition at F1 of 0.62. These weaknesses propagated to downstream relation extraction, where the best ADE-drug system achieved only F1 of 0.43, and the best Reason-drug system achieved F1 of 0.50.

In a study that aimed at assessing the generalizability of NLP-based machine learning models for detecting Drug-ADE relationships across clinical corpora [10]. The authors first constructed an institution-specific dataset linking drugs to ADEs using records from The Ohio State University James Cancer Hospital. Then they used the N2C2 2018 dataset as an external benchmark to test cross-domain transfer. The authors compared a range of approaches, which include SVMs, CNNs, BiLSTMs, and transformer-based models (BERT and ClinicalBERT), by training on one corpus and testing on the other. Across both transfer directions, ClinicalBERT achieved the strongest performance, obtaining an F1 of 0.78 when trained on the constructed dataset and evaluated on N2C2 (vs. 0.61-0.73 for other methods), and an F1 of 0.74 when trained on N2C2 and evaluated on the constructed dataset (vs. 0.55-0.72 for other methods). Importantly, their experiments focus on relation classification using pre-specified relation instances rather than a complete end-to-end setting; that is, the evaluation assumes access to the relevant entities/pairs and does not include the upstream entity detection step that typically drives error propagation in end-to-end extraction.

Another study used the N2C2 2018 dataset to explore the relationship between a drug and its associated entities using relation extraction methods [11]. In this study, the authors used three approaches to complete that task: a rule-based approach, a deep learning-based approach, and a contextualized language model-based approach. For the rule-based approach, a breadth-first search algorithm was used to find the nearest occurrence of the drug on either side of the non-drug entity. As for the deep learning approach, CNNs were used, incorporating four primary layers: embedding, convolution, pooling, and feed-forward layers, with pre-trained word vectors from word2vec and GloVe as inputs. The task is approached as a binary classification problem, creating separate models for each drug-entity pair. Finally, for the contextualized language model-based approach, BERT (cased and uncased), BioBERT, and ClinicalBERT were used. Results demonstrated that the approach based on contextualized language models surpassed all other models in performance, achieving the highest standard in ADE extraction with a Precision of 0.93, a Recall of 0.96, and an F1 score of 0.94. Nonetheless, for specific types of relations, the rule-based method demonstrated superior Precision and Recall compared to any of the learning-based.

[12] Focused on concept extraction of ADEs and medication entities, the aim was to evaluate several biomedical contextual embeddings, including BERT, ELMo, and Flair. The N2C2 2018 dataset is also used in that study. The study explores best practices for transfer learning, such as fine-tuning language models and employing scalar mix techniques, achieving overall task performance (F1=0.93) and specifically in ADE identification (F1=0.53). It finds that Flair-based embeddings excel at recognizing context-dependent entities such as ADEs, while BERT-based embeddings are superior at identifying clinical terminology, including Drug and Form entities. Furthermore, they developed a sentence augmentation technique to enhance ADE extraction and reported ADE extraction performance of (F1= 0.55).

## III. The N2C2 Dataset & Preliminary findings

### A. The N2C2 2018 Dataset

The dataset we focus on and use is the N2C2 dataset from the 2018 National NLP Clinical Challenges shared task track 2. Track 2 challenges focused on extracting ADEs from clinical notes, with main tasks including concept extraction, relation classification, and end-to-end systems [8]. The dataset contains 505 annotated notes: 303 for training and 202 for testing.

The first subtask of Track 2 is the concept extraction task, which focuses on identifying predefined types of clinical entities from the clinical notes. Two of these types are ADE and Reason. The next subtask is the relation extraction task, where the systems are provided with a gold-standard set of annotated entities and are required to classify the relations (including ADE and Reason as relations) between them. The end-to-end task evaluates systems on raw, unannotated clinical text, requiring joint entity detection and relation classification. Therefore, errors introduced during the concept extraction phase directly propagate to the relation extraction stage, as entities that are not detected cannot participate in downstream relation classification. We hypothesize that end-to-end performance is largely constrained by this pipeline dependency, with overall results primarily limited by the upstream entity detection step.

### B. Preliminary Findings

First, we conducted a preliminary analysis to identify the challenges. We followed the same task formulation as the N2C2 2018 shared task and trained a BioMedBERT-BiLSTM-CRF model for the concept extraction stage. The model achieved strong performance on drug mentions but substantially lower performance on ADE and Reason. Specifically, the model achieved 0.95 precision, 0.96 recall, and an F1 of 0.958 for Drugs, matching prior high performance results on the N2C2 2018 dataset [8]. On the other hand, performance for Reasons (F1 of 0.72) and ADEs (F1 of 0.60) remained much lower, though it improved slightly over prior benchmarks for ADE. This pattern is consistent with the broader literature [8], [12], where even state-of-the-art systems report comparatively limited accuracy for ADE and Reason extraction. Overall, these results reinforce that ADE and Reason mentions remain the main bottleneck for end-to-end drug-event relation extraction and downstream pharmacovigilance applications.

#### 1) What is annotated as ADE or as a Reason

Here, we provide examples that illustrate why extracting ADE and REASON as entities prior to Relation Classification can be challenging. Adverse-event mentions can appear in multiple forms, including signs or symptoms (e.g., “rash”), diseases or conditions (e.g., “renal failure”), and measurement-based statements such as abnormal lab or vital values (e.g., “elevated BP” or “increased INR”). Furthermore, a symptom (e.g., rash) can appear as an ADE when “cefepime was discontinued secondary to drug rash,” while a similar symptom may be a treatment target in a Reason context, such as “Started miconazole… for rash.” Likewise, a condition may be described as drug-related harm (“renal failure… secondary to CSA”) or as the motivation for therapy (“renal failure worsened and was switched to IV vancomycin”). Additionally, the same AE may also be documented as part of the patient’s baseline history or ongoing disease course with no medication linkage; in such cases, it is not an ADE or a Reason. Thus, under the N2C2 2018 annotation scheme, such mentions would typically not be annotated because they do not participate in a drug-event relation [8].

### C. Challenges and Design Alternatives

Prior work on the N2C2 2018 benchmark shows that concept extraction models perform well on medications and structured attributes, but consistently struggle to detect ADE and Reason entities, making these event types a key source of error [8], [12]. Following the standard shared-task formulation, we first attempted to detect entities as distinct types, including ADE and Reason. As illustrated above, the same term (e.g., rash) can correspond to either an ADE or a Reason, and the correct label can be determined only by considering the surrounding context that specifies the event’s relationship to the drug. However, the relation context, the text linking the term to the drug, is considered in the next step, relation extraction.

We believe that current concept extraction systems consistently underperform in identifying entity types. A major contributor to this gap is the widespread practice of treating “ADEs” and “Reasons” as distinct concept labels during training. This practice forces models to learn separate decisions for phenomena that are clinically identical except for their pharmacological relationship to a drug. The resulting low concept-extraction results for AEs harm the relation-extraction task, because these entities were not detected in the first step.

Therefore, to advance the extraction of ADEs, we can adopt either of two complementary strategies that can address this bottleneck: (i) jointly modeling concept extraction and relation extraction, or (ii) defining a unified, more general event concept that covers both ADE and Reason mentions during extraction and then deferring causal interpretation to a dedicated relation classification layer. We chose the second approach because separating concept extraction from relation classification simplifies both tasks and aligns with prior evidence that relation classification can be performed reliably when candidate pairs are available [8]. An overview of the proposed pipeline is provided in Fig 1.

**Fig. 1.**
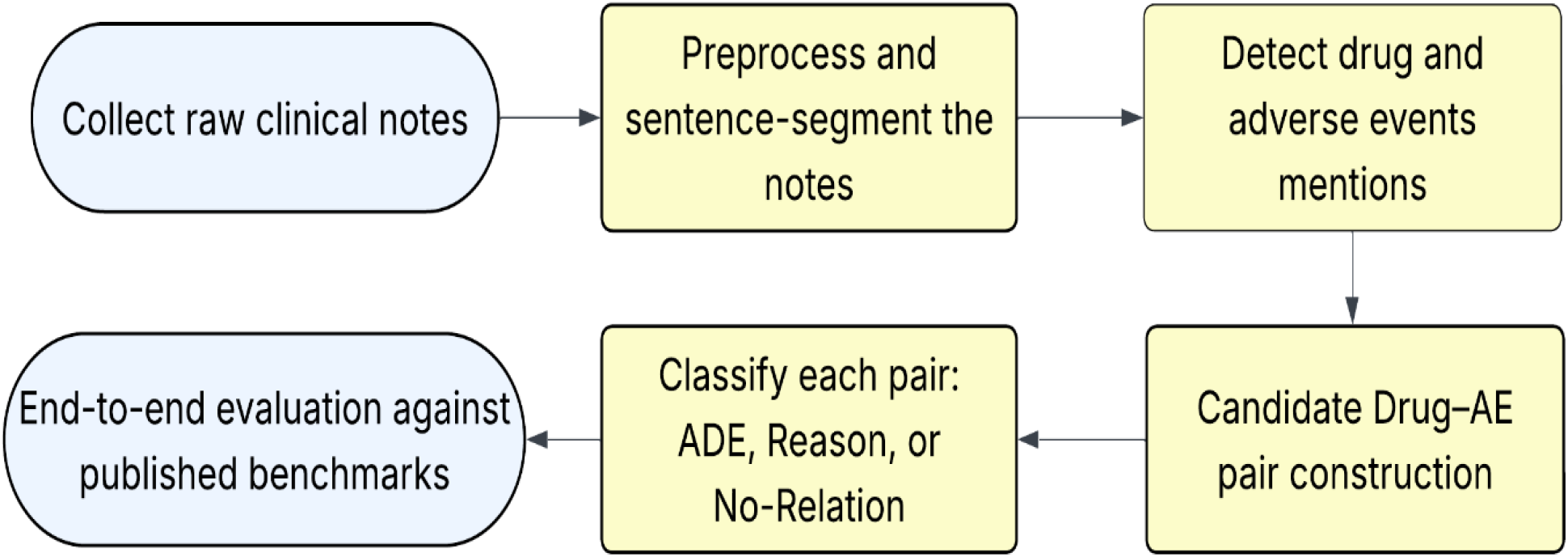
Overview of the proposed end-to-end pipeline for drug-event extraction from clinical notes, including sentence segmentation, drug and AE mention detection, candidate Drug-AE pair generation, relation classification (Drug-ADE, Drug-Reason, No-Relation), and finaly, evaluation

## IV. Methods

### A. Drug and AE detection

Building on the error propagation observed in the end-to-end setting, we developed an extraction pipeline designed to improve Drug-ADE and Drug-Reason identification in clinical notes. We first detect drug mentions using our BioMedBERT-BiLSTM-CRF concept extraction model. Instead of differentiating ADE versus Reason at the concept extraction stage, we defer this distinction to the relation classification stage. Thus, during concept extraction, we treat both ADE and Reason mentions as a single unified event type, which we refer to as an adverse event (AE). Which raises a practical question: how can we train an AE detector? Existing datasets lack annotations for a concept that generalizes over ADE and Reason entities. Moreover, the N2C2 annotations cannot be directly repurposed because they label only events that participate in Drug-ADE or Drug-Reason relations; clinically relevant events not linked to a drug are typically unannotated [8]. Therefore, these unmarked event mentions cannot be treated as true negatives for AE detection.

To support robust AE detection, we explored two complementary strategies: (1) adapting an existing clinical concept extraction system for AE identification and (2) using an LLM for adverse-event span detection. For the first strategy, we used the Medical NER system to identify clinical event mentions from narrative text [13]. For the second strategy, we used an OpenAI GPT model to identify AE mentions in clinical notes [14].

#### 1) Medical NER (BERT-based)

We leveraged an existing transformer-based clinical NER tool, blaze999/Medical-NER, as a general-purpose concept extractor for broad event detection [13]. This model is a DeBERTa-based token classifier trained to recognize 41 medical entity types, but we focused on a subset of labels that most directly aligned with AE mentions. Specifically, we extracted spans predicted as ‘Sign Symptom’ and ‘Disease Disorder’ as AE candidates, since these categories cover a large portion of the AEs that later participate in Drug-ADE and Drug-Reason relations. Additionally, we increased coverage using rule-based patterns to capture AE-relevant expressions that this tool did not capture (e.g., abnormal lab or vital-sign values).

#### 2) GPT model (LLM-based)

For the LLM-based approach, we used GPT-4o-mini as an adverse-event span detector operating directly over sentence-level clinical text [14]. Each sentence was provided to the model with a fixed prompt instructing it to act as a “precise clinical NER highlighter” and to extract all spans corresponding to AEs, defined broadly to include symptoms, medical conditions, diseases/disorders, infections, abnormal laboratory results, and abnormal vital signs. See Appendix A for the full prompt.

### B. Relation Extraction

To construct the relation extraction (RE) dataset, we generated candidate drug-event pairs at the sentence level. For each sentence containing N detected drugs and M AE candidates, we created N × M Drug-AE pairs by inserting [D]…[/D] around the drug mention and [AE]…[/AE] around the candidate event span while preserving the original sentence text. Each pair was then assigned one of three labels, Drug-ADE, Drug-Reason, or No-Relation, by aligning the candidate event span to the gold annotations of the N2C2 dataset and verifying whether the paired drug matches the drug linked to that event in the annotated relation. Pairs whose event candidate overlapped a gold ADE span and was linked to the same drug were labeled Drug-ADE; pairs overlapping a gold Reason span and linked to the same drug were labeled Drug-Reason; all remaining combinations were labeled No-Relation.

As for relation classification, we used T5-based sequence-to-sequence models adapted for clinical text [15]. These models were pre-trained on large biomedical and clinical corpora, including MIMIC-IV and PubMed Central (PMC), and leverage MIMIC-informed vocabulary to represent domain-specific terminology for clinical relation classification better.

### C. End-to-end evaluation

Figure 1 summarizes the end-to-end setting, in which the system operates on raw, unannotated clinical notes and must perform the full extraction workflow without access to gold entities. After preprocessing and sentence segmentation, the pipeline detects drug mentions and AE mentions directly from text, constructs candidate Drug-AE pairs by combining each detected drug with each detected AE, and then classifies each pair as Drug-ADE, Drug-Reason, or No-Relation. This setting reflects real-world deployment because upstream mention-detection errors directly determine which relations can be formed and evaluated downstream.

We report performance using precision, recall, and F1-score, and we report lenient F1 to match the official N2C2 2018 evaluation protocol. Finally, we benchmark our end-to-end results against previously reported state-of-the-art end-to-end systems on the same task to contextualize the impact of our AE detection and pairing strategies [8].

## V. Results

### A. Relation Extraction

In the relation classification setting, where gold-standard drug-event pairs were provided, and the model was only tasked with classifying the relation type, the T5-EHR model achieved consistently strong performance, see Table III. It reached an F1 of 0.98 for Drug-Reason relations and an F1 of 0.94 for Drug-ADE relations. These results align with prior n2c2 2018 relation classification studies, which report similarly high performance when entity pairs are provided; for example, [16] reports an F1 of 0.97 for both Drug-ADE and Drug-Reason relations using BERT-family models. Thus, results indicate that, once entity pairs are identified, distinguishing whether a drug has caused an adverse event versus a therapeutic indication is highly learnable. This finding supports the broader conclusion that end-to-end performance is often limited less by relation classification itself and more by upstream event detection and pairing, emphasizing that the overall pipeline design and concept extraction strategy are key to improving end-to-end drug-event relation extraction.

**TABLE I.** Statistics for concept extraction in the n2c2 2018 dataset.

| Entities | Concept Extraction Samples |  |  |
| --- | --- | --- | --- |
|  | Full | Train | Test |
| Drug | 26,800 | 16,225 | 10,757 |
| Strength | 10,921 | 6,691 | 4,230 |
| Form | 11,010 | 6,651 | 4,359 |
| Dosage | 6,902 | 4,221 | 2,681 |
| Frequency | 10,293 | 6,281 | 4,012 |
| Route | 8,989 | 5,476 | 3,513 |
| Duration | 970 | 592 | 378 |
| Reason | 6,400 | 3,855 | 2,545 |
| ADE | 1,584 | 959 | 625 |
| Total | 83,869 | 50,951 | 32,918 |

**TABLE II.** Statistics for the relation identification in the n2c2 2018 dataset.

| Relations | Relations Identification Samples |  |  |
| --- | --- | --- | --- |
|  | Full | Train | Test |
| Strength-Drug | 10,946 | 6,702 | 4,244 |
| Form-Drug | 11,028 | 6,654 | 4,374 |
| Dosage-Drug | 6,920 | 4,225 | 2,695 |
| Frequency-Drug | 10,344 | 6,310 | 4,034 |
| Route-Drug | 9,084 | 5,538 | 3,546 |
| Duration-Drug | 1,069 | 643 | 426 |
| Reason-Drug | 8,579 | 5,169 | 3,410 |
| ADE-Drug | 1,840 | 1,107 | 733 |
| Total | 59,810 | 36,384 | 23,462 |

**TABLE III.**
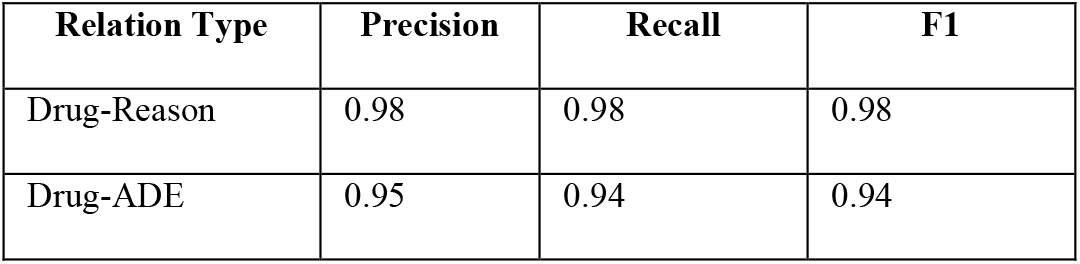
t5-ehr performance on relation classification with gold drug-event pairs (not end-to-end)

### B. End to-end Results

In the end-to-end relation extraction setting, we evaluated performance separately for Drug-Reason and Drug-ADE by combining each AE-detection strategy with a Clinical-T5 relation classifier. Using LLM-derived AE annotations, T5-EHR achieved 0.98 precision, 0.89 recall, and 0.94 F1 for Drug-Reason, and 0.94 precision, 0.91 recall, and 0.93 F1 for Drug-ADE, see Table IV. With Medical NER-derived AE annotations, performance remained strong but was consistently lower, reaching 0.90 F1 for Drug-Reason (0.97 precision; 0.82 recall) and 0.91 F1 for Drug-ADE (0.94 precision; 0.88 recall). Both T5-EHR configurations substantially outperformed previously reported state-of-the-art end-to-end benchmarks (F1 of 0.59 for Drug-Reason and 0.48 for Drug-ADE), with the LLM-based AE detection yielding the best overall end-to-end results.

**TABLE IV.** end-to-end drug-event extraction results on n2c2 2018 using ae detection + t5-ehr relation classification (lenient f1):

| AE detection method | Classification model | Relation Type | Precision | Recall | F1 |
| --- | --- | --- | --- | --- | --- |
| LLM (GPT-4o-mini) | T5-EHR | Drug-Reason | 0.98 | 0.89 | 0.94 |
|  |  | Drug-ADE | 0.94 | 0.91 | 0.93 |
| Medical NER | T5-EHR | Drug-Reason | 0.97 | 0.82 | 0.90 |
|  |  | Drug-ADE | 0.94 | 0.88 | 0.91 |
| State-of-the-art [8] |  | Drug-Reason | - | - | 0.59 |
|  |  | Drug-ADE | - | - | 0.48 |

### C. End to-end Results Using Only the LLM

In addition, we evaluated an alternative LLM-only end-to-end configuration. Since the LLM performed well at zero-shot AE detection, we also tested whether it could carry the pipeline end-to-end by performing relation classification directly after AE detection. Under this setting, the LLM achieved F1 = 0.78 for Drug-Reason and F1 = 0.67 for Drug-ADE, as shown in Table V [8]. These results indicate that, while an LLM can provide a reasonable zero-shot baseline, supervised training remains important for accurate relation classification, consistent with the substantially higher performance achieved by the trained T5-EHR relation model [15].

**TABLE V.** end-to-end results using an llm-only pipeline (gpt-4o mini) for ae detection and relation classification:

| AE detection method | Classification model | Relation Type | Precision | Recall | F1 |
| --- | --- | --- | --- | --- | --- |
| LLM (GPT-4o-mini) | LLM (GPT-4o-mini) | Drug-Reason | 0.68 | 0.93 | 0.78 |
|  |  | Drug-ADE | 0.53 | 0.90 | 0.67 |

## VI. Discussion

Our initial findings showed that the BioMedBERT-BiLSTM-CRF architecture is highly accurate for structured entities; however, concept extraction of ADE and Reason entities remains significantly lower. While these predictions support strong performance on template-like relations such as Drug-Strength, Drug-Form, and Drug-Duration, end-to-end extraction performance remains much lower for the clinically critical relations Drug-ADE and Drug-Reason. With the intention of distinguishing between ADE and Reason at the relation classification level rather than concept extraction, we propose a new approach. In this approach, we first recognize a new type, Adverse Events (AE), and hoping to detect all AEs in the clinical narrative as comprehensively as possible, independent of whether they are ultimately linked to a drug, we then use relation classification to see if the AEs participate in Drug-AE relation, assigning each Drug-event pair to Drug-ADE, Drug-Reason, or No-Relation.

The N2C2 2018 dataset also introduces a second structural challenge, that is, partial labeling of AEs. Many AE mentions are not annotated in N2C2 when they do not participate in an annotated Drug-ADE or Drug-Reason relation [8]. Because these unlabeled AE-like mentions vastly outnumber labeled ADE and Reason instances, the model is repeatedly exposed to the same clinical concepts labeled as ‘no type’ far more often than as ADE or Reason. This leads to poor detection of AEs, even when they represent ADE or Reason cases, undermining end-to-end performance.

To mitigate these limitations and reduce error propagation in the end-to-end setting, we introduced an AE detection and relation construction pipeline and compared two AE detection strategies: Medical NER and LLM-based span detection [13], [14]. Both strategies increased AE coverage compared with the base concept extraction model, but the LLM approach missed fewer gold events and produced more exact span matches. This matters because end-to-end relation extraction is more reliable when AE spans are complete and precisely bounded, since accurate boundaries improve both drug-event pairing and consistent relation labeling.

These improvements in AE detection translated into significantly stronger end-to-end relation performance. Using the same T5-based relation classifier [15], both pipelines substantially exceeded previously reported end-to-end benchmarks, with the LLM-based AE detection providing the best overall results as reported in Table IV. This outcome indicates that end-to-end performance in the N2C2 setting depends more on accurately detecting events and capturing their whole spans than on the relation classifier itself. Put simply, improving AE candidate detection removes the main bottleneck and allows the relation model to perform closer to its potential.

## VII. Conclusion

We have reformulated our approach to high-recall, high-quality ADEs for pharmacovigilance. This change was driven by the recognition that dedicated AE datasets annotated independently of drug relations are necessary for more reliable end-to-end extraction. Since we are not aware of any dataset for training the recognition of adverse events from clinical text, we explored adapting two existing resources for AE recognition. Combined with the use of the relation classification model, we substantially improve the N2C2 2018 Track 2 end-to-end results compared with prior benchmarks.

## VIII. Future directions

Our findings strongly suggest the need for a dedicated dataset that annotates adverse events in clinical notes, regardless of whether they are drug-related. Current benchmarks, primarily the N2C2 2018 [8], annotate adverse event mentions only when they participate in a drug-event relation, which limits the development and evaluation of robust AE detectors and complicates end-to-end extraction.

To address this gap, our future work will focus on creating an AE-focused benchmark that mirrors the n2c2 2018 evaluation style while removing the dependency between mention annotation and drug linkage. Specifically, we will create a corpus annotated for Drug and AE mentions at the entity level, and then provide three aligned evaluation tracks:

1. Concept Extraction, where systems detect Drugs and AEs from raw text.
2. Relation Extraction, where gold Drug-AE pairs are provided and systems classify relations (e.g., Drug-ADE, Drug-Reason, No-Relation).
3. End-to-End Evaluation, where systems must jointly detect entities and classify drug-event relations from unannotated notes.

This design will enable more reliable measurement of AE detection quality, clearer separation of error sources across pipeline stages, and stronger end-to-end pharmacovigilance systems.

## Data Availability

Access to the dataset is available through the official N2C2 data request process and requires completion of the applicable data-use agreement and credentialing requirements.

https://n2c2.dbmi.hms.harvard.edu/2018-challenge

## Appendix

### A. LLM Prompt for AE Span Detection

“*You are a precise clinical NER highlighter. I will give you a set of sentences. For each sentence, identify all spans of one or more tokens that are ‘Adverse Events,’ where an adverse event is a symptom, medical condition, disease, disorder, bacterial or viral infection, abnormal lab result, or abnormal vital sign. Sentences:”*

### B. Parameter Choices

- **Environment**: Google Colab
- **Computational resources:** TPU (T5-EHR), NVIDIA A100 GPU (BioMedBERT-BiLSTM-CRF)
- **T5-EHR parameters:**
  - 10 epochs; LR = 1e−4; warmup = 0
  - Batch size: train = 1; eval = 32
  - Sequence lengths: max input = 256
- **BioMedBERT-BiLSTM-CRF parameters:**
  - 10 epochs; LR = 2e−5; warmup = 0
  - Batch size: train = 32; eval = 32
  - Sequence lengths: max input = 256

## Acknowledgment

We acknowledge financial support for O.A. from the Saudi Arabian Cultural Mission (SACM) throughout his Ph.D. studies. C.C. was partially funded by an Institutional Development Award (IDeA) from the National Institute of General Medical Sciences of the National Institutes of Health under awards numbers U54GM104941 and P20GM103446.

## Notes

### Competing Interest Statement

The authors have declared no competing interest.

### Author Declarations

This study used de-identified clinical data from the 2018 n2c2/National NLP Clinical Challenges shared task dataset, which is derived from clinical notes released under controlled-access data-use conditions. Access was obtained through the required N2C2 Data Use Agreement administered by the Harvard Medical School Department of Biomedical Informatics. No attempt was made to identify individual patients. The original MIMIC-III database was approved by the Institutional Review Boards of Beth Israel Deaconess Medical Center and Massachusetts Institute of Technology, with waiver of individual patient consent because the data were de-identified.

## References

[1] World Health Organization (WHO). International drug monitoring: the role of national centres. WHO Technical Report Series 1972;498.

[2] Kommu S., Carter C., and Whitfield P., “Adverse Drug Reactions,” in StatPearls [Internet]. Treasure Island (FL): StatPearls Publishing, updated Jan. 10, 2024. [Online]. Available: https://www.ncbi.nlm.nih.gov/books/NBK599521/.

[3] Centers for Disease Control and Prevention, “FastStats: Medication Safety Data,” CDC, Apr. 17, 2024. [Online]. Available: https://www.cdc.gov/medication-safety/data-research/facts-stats/index.html.

[4] W. Wang, D. Ferrari, G. Haddon-Hill, et al., “Electronic health records as source of research data,” in Machine Learning for Brain Disorders, O. Colliot, Ed. Humana, 2023, ch. 11, doi: 10.1007/978-1-0716-3195-9_11.

[5] M. K. Kim, C. Rouphael, J. McMichael, N. Welch, and S. Dasarathy, “Challenges in and opportunities for electronic health record-based data analysis and interpretation,” Gut and Liver, vol. 18, no. 2, pp. 201–208, 2024, doi: 10.5009/gnl230272.

[6] S. Modi, K. A. Kasmiran, N. Mohd Sharef, and M. Y. Sharum, “Extracting adverse drug events from clinical notes: A systematic review of approaches used,” Journal of Biomedical Informatics, vol. 151, Art. no. 104603, 2024, doi: 10.1016/j.jbi.2024.104603.

[7] S. Kim, T. Kang, T. K. Chung, Y. Choi, Y. Hong, K. Jung, and H. Lee, “Automatic extraction of comprehensive drug safety information from adverse drug event narratives in the Korea Adverse Event Reporting System using natural language processing techniques,” Drug Safety, vol. 46, no. 8, pp. 781–795, 2023, doi: 10.1007/s40264-023-01323-2.

[8] S. Henry, K. Buchan, M. Filannino, A. Stubbs, and O. Uzuner, “2018 n2c2 shared task on adverse drug events and medication extraction in electronic health records,” Journal of the American Medical Informatics Association, vol. 27, no. 1, pp. 3–12, 2020, doi: 10.1093/jamia/ocz166

[9] A. Jagannatha, F. Liu, W. Liu, and H. Yu, “Overview of the first natural language processing challenge for extracting medication, indication, and adverse drug events from electronic health record notes (MADE 1.0),” Drug Safety, vol. 42, no. 1, pp. 99–111, 2019, doi: 10.1007/s40264-018-0762-z.

[10] M. M. Zitu, S. Zhang, D. H. Owen, C. Chiang, and L. Li, “Generalizability of machine learning methods in detecting adverse drug events from clinical narratives in electronic medical records,” Frontiers in Pharmacology, vol. 14, Art. no. 1218679, 2023, doi: 10.3389/fphar.2023.1218679.

[11] D. Mahendran and B. T. McInnes, “Extracting adverse drug events from clinical notes,” in AMIA Joint Summits on Translational Science Proceedings, 2021, pp. 420–429.

[12] S. Narayanan, K. Mannam, S. P. Rajan, and P. V. Rangan, “Evaluation of transfer learning for adverse drug event (ADE) and medication entity extraction,” in Proc. 3rd Clinical Natural Language Processing Workshop, Online, Nov. 2020, pp. 55–64, doi: 10.18653/v1/2020.clinicalnlp-1.6

[13] S. Mattupalli, “Medical-NER” [Pretrained model], Hugging Face, 2023. [Online]. Available: https://huggingface.co/blaze999/Medical-NER

[14] OpenAI, “GPT-4o mini: advancing cost-efficient intelligence,” Jul. 18, 2024. [Online]. Available: https://openai.com/index/gpt-4o-mini-advancing-cost-efficient-intelligence/

[15] S. M. S. Althabiti, “Developing generative language models for clinical natural language processing,” Ph.D. dissertation, University of Delaware, Newark, DE, USA, 2025. [Online]. Available: https://www.proquest.com/dissertations-theses/developing-generative-language-models-clinical/docview/3290574087/se-2

[16] D. Mahendran and B. T. McInnes, “Extracting adverse drug events from clinical notes,” AMIA Joint Summits on Translational Science Proceedings, vol. 2021, pp. 420–429, May 2021. PMID: 34457157. PMCID: PMC8378605

